# Testing the association between blood type and COVID-19 infection, intubation, and death

**DOI:** 10.1101/2020.04.08.20058073

**Authors:** Michael Zietz, Jason Zucker, Nicholas P. Tatonetti

**Author notes:** Corresponding author (Nicholas P. Tatonetti).

## Abstract

The rapid global spread of the novel coronavirus SARS-CoV-2 has strained healthcare and testing resources, making the identification and prioritization of individuals most at-risk a critical challenge. Recent evidence suggests blood type may affect risk of severe COVID-19. We used observational healthcare data on 14,112 individuals tested for SARS-CoV-2 with known blood type in the New York Presbyterian (NYP) hospital system to assess the association between ABO and Rh blood types and infection, intubation, and death. We found slightly increased infection prevalence among non-O types. Risk of intubation was decreased among A and increased among AB and B types, compared with type O, while risk of death was increased for type AB and decreased for types A and B. We estimated Rh-negative blood type to have a protective effect for all three outcomes. Our results add to the growing body of evidence suggesting blood type may play a role in COVID-19.

## Background

The novel Coronavirus disease (COVID-19, caused by the SARS-CoV-2 virus) has spread rapidly across the globe and has caused over 21.1 million confirmed infections and over 761,000 deaths worldwide as of August 17, 2020 [1]. A number of risk factors for COVID-19 morbidity and mortality are known, including age, sex, smoking, hypertension, diabetes, and chronic cardiovascular and respiratory diseases [2,3].

Recent work has demonstrated an association between ABO blood types and COVID-19 risk. Using data from Wuhan and Shenzhen, Zhao et al. found a greater proportion of A and a lower proportion of O blood types among COVID-19 patients, relative to the general populations of Wuhan and Shenzhen [4]. Similarly, using a meta-analysis of data from Italy and Spain, Ellinghaus et al. [5] found a higher risk of COVID-19 among A and a lower risk among O blood types. Conversely, however, they estimated lower odds of mechanical ventilation for all non-O types, though the estimated odds ratios were not statistically significant at the 5% level for this outcome.

The ABO blood type trait reflects polymorphisms within the *ABO* gene. This gene is associated with a number of other traits, including risk factors for COVID-19 morbidity and mortality. For example, genome-wide association studies have associated variants within *ABO* to activity of the angiotensin converting enzyme (ACE) [6], red blood cell count, hemoglobin concentration, hematocrit, [7,8,9,10], von Willebrand factor [11,12,13,14], myocardial infarction [15,16], coronary artery disease [16,17,18,19,20], ischemic stroke [12,18,21], type 2 diabetes [22,23,24], and venous thromboembolism [25,26,27,28,29,30,31,32]. A 2012 meta-analysis found that, in addition to individual variants, a non-O blood type is among the most important genetic risk factors for venous thromboembolism [33]. These conditions are also relevant for COVID-19. For example, coagulopathy is a common issue for COVID-19 patients [34,35,36,37,38,39,40], and risk of venous thromboembolism must be carefully managed [41].

The numerous associations between conditions and both blood type and COVID-19 provide reason to believe true associations may exist between blood type and morbidity and mortality due to COVID-19. In addition, previous work has identified associations between ABO blood groups and a number of different infections or disease severity following infections, including SARS-CoV-1 [42], *P. falciparum* [43], *H. pylori* [44], Norwalk virus [45], hepatitis B virus [46], and *N. gonorrhoeae* [47].

Rh(D) phenotypes (positive and negative Rh blood types) are associated with very few diseases compared to ABO [48]. Like ABO, Rh type is important for type compatibility and immune response. For example, hemolytic disease of the newborn is a concern when Rh(D) is mismatched between mother and offspring [49]. Other studies have found evidence that Rh-positive individuals are protected against the effects of latent toxoplasmosis [50], though *Toxoplasma gondii* is a eukaryotic parasite [51], not a virus like SARS-CoV-2.

Within the United States, New York suffered among the worst outbreaks during the early phases of the pandemic. As of August 22, New York City has recorded 228,144 confirmed infections and 19,014 deaths [52]. We sought to understand the association between SARS-CoV-2 infection/COVID-19 and blood type using electronic health record (EHR) data from New York-Presbyterian/Columbia University Irving Medical Center (NYP/CUIMC) hospital in New York City, USA. We compared both ABO and Rh(D) blood types, and we investigated initial infection status and two severe COVID-19 outcomes: intubation and death. We evaluated potential confounding due to population stratification using a multivariate analysis, and we report clinically meaningful measures of effect.

## Results

We determined blood types using laboratory measurements recorded in the NYP/CUIMC EHR system. After removing likely errors, such as individuals with contradictory blood type results, we identified 14,112 adult individuals with known blood types who received at least one SARS-CoV-2 swab test (Table 1, Supplementary Figure S1). We performed chi-squared tests of independence and found insufficient evidence to conclude that the blood group frequencies differ between SARS-CoV-2-tested and non-tested groups (Supplementary Table 1). Individuals were considered initially SARS-CoV-2 positive (COV+) if they tested positive on their first recorded test or within the following 96 hours. We evaluated associations between blood types and outcomes using three comparisons: infection prevalence among initial tests and survival analysis for intubation and death among individuals with infections confirmed by swab test. We report on clinical data as of August 1, 2020.

**Table 1:** Summary demographics for SARS-CoV-2-tested individuals at NYP/CUIMC, stratified by blood type. N is the number of individuals having the given blood type who had at least one recorded test for SARS-CoV-2. Age is reported as the median and interquartile range (25-75). Percents are reported relative to individuals having the blood type, except the N row, where percents are by blood group type (ABO or Rh) and are relative to all individuals in the study.

|  | **A** | **AB** | **B** | **O** | **Rh-neg** | **Rh-pos** |
| --- | --- | --- | --- | --- | --- | --- |
| N | 4298 (32.9) | 559 (4.3) | 2033 (15.6) | 6161 (47.2) | 1195 (9.2) | 11856 (90.8) |
| Age (IQR) | 58 (37 to 72) | 57 (37 to 71) | 57 (37 to 72) | 55 (36 to 71) | 56 (37 to 70) | 56 (37 to 71) |
| Male (%) | 1676 (39.0) | 231 (41.3) | 778 (38.3) | 2339 (38.0) | 430 (36.0) | 4594 (38.7) |
| Hispanic (%) | 1572 (36.6) | 173 (30.9) | 666 (32.8) | 2583 (41.9) | 389 (32.6) | 4605 (38.8) |
| **Race** |  |  |  |  |  |  |
| Asian (%) | 71 (1.7) | 21 (3.8) | 89 (4.4) | 123 (2.0) | 16 (1.3) | 288 (2.4) |
| Black (%) | 629 (14.6) | 95 (17.0) | 493 (24.2) | 1179 (19.1) | 151 (12.6) | 2245 (18.9) |
| Missing (%) | 728 (16.9) | 79 (14.1) | 370 (18.2) | 1093 (17.7) | 192 (16.1) | 2078 (17.5) |
| Other (%) | 1085 (25.2) | 132 (23.6) | 464 (22.8) | 1715 (27.8) | 263 (22.0) | 3133 (26.4) |
| White (%) | 1785 (41.5) | 232 (41.5) | 617 (30.3) | 2051 (33.3) | 573 (47.9) | 4112 (34.7) |
| **Outcomes** |  |  |  |  |  |  |
| Initially COV+ (%) | 754 (17.5) | 88 (15.7) | 363 (17.9) | 1060 (17.2) | 164 (13.7) | 2101 (17.7) |
| COV+ (%) | 786 (18.3) | 94 (16.8) | 392 (19.3) | 1122 (18.2) | 175 (14.6) | 2219 (18.7) |
| COV+/Intubated (%) | 111 (2.6) | 17 (3.0) | 78 (3.8) | 193 (3.1) | 24 (2.0) | 375 (3.2) |
| COV+/Died (%) | 104 (2.4) | 15 (2.7) | 46 (2.3) | 166 (2.7) | 11 (0.9) | 320 (2.7) |

The unadjusted prevalence of initial infection was higher among A and B blood types and lower among AB types, compared with type O (Table 2, Figure 1). To avoid bias with respect to healthcare utilization, length of hospital stay, and potential in-patient infection, we evaluated the prevalence of infection among individuals only during the first encounters in which they were tested. In addition, to account for the considerable risk of false negative tests [55,56] and the fact that providers would repeat the test in patients with high clinical suspicion for COVID-19 [57], any positive test during the first 96 hours of an encounter was considered evidence of initial infection.

**Table 2:** Effect size estimates for blood types with and without correction for race and ethnicity. Risks computed using linear regression (for prevalence) or the cumulative incidence from Fine-Gray models (for intubation and death). Risk differences and ratios computed relative to O ABO blood type and positive Rh(D) type.

| Outcome | Blood type | Unadjusted |  |  | Race/ethnicity adjusted |  |  |
| --- | --- | --- | --- | --- | --- | --- | --- |
|  |  | Risk | Risk difference | Risk ratio | Risk | Risk difference | Risk ratio |
| Prevalence | A | 17.5 (16.3 to 18.8) | 0.3 (-1.2 to 1.9) | 1.02 (0.93 to 1.12) | 18.0 (16.8 to 19.2) | 1.3 (-0.3 to 3.0) | 1.08 (0.98 to 1.19) |
|  | AB | 15.7 (12.8 to 18.7) | -1.5 (-4.5 to 1.7) | 0.91 (0.74 to 1.10) | 16.8 (13.9 to 19.8) | 0.1 (-2.8 to 3.2) | 1.01 (0.83 to 1.20) |
|  | B | 17.9 (16.1 to 19.5) | 0.7 (-1.4 to 2.6) | 1.04 (0.92 to 1.16) | 18.0 (16.3 to 19.7) | 1.3 (-0.7 to 3.3) | 1.08 (0.96 to 1.20) |
|  | O | 17.2 (16.2 to 18.1) | — | — | 16.7 (15.7 to 17.6) | — | — |
|  | Rh-pos | 17.7 (17.0 to 18.4) | — | — | 17.6 (16.9 to 18.3) | — | — |
|  | Rh-neg | 13.7 (11.8 to 15.5) | -4.0 (-6.1 to -2.0) | 0.77 (0.66 to 0.88) | 14.9 (13.0 to 16.8) | -2.7 (-4.7 to -0.8) | 0.85 (0.73 to 0.96) |
| Intubation | A | 17.2 (14.2 to 20.1) | -3.2 (-7.5 to 0.3) | 0.84 (0.66 to 1.02) | 17.3 (14.3 to 20.4) | -2.9 (-7.2 to 0.6) | 0.85 (0.68 to 1.03) |
|  | AB | 21.8 (12.3 to 31.7) | 1.4 (-8.5 to 11.6) | 1.07 (0.59 to 1.57) | 22.1 (12.8 to 32.1) | 1.8 (-8.3 to 12.2) | 1.09 (0.60 to 1.59) |
|  | B | 22.9 (18.6 to 27.6) | 2.5 (-2.5 to 7.6) | 1.12 (0.89 to 1.40) | 22.8 (18.6 to 27.5) | 2.5 (-2.7 to 7.5) | 1.12 (0.88 to 1.40) |
|  | O | 20.4 (17.8 to 23.4) | — | — | 20.3 (17.7 to 23.3) | — | — |
|  | Rh-pos | 20.3 (18.4 to 22.1) | — | — | 20.2 (18.4 to 22.1) | — | — |
|  | Rh-neg | 14.6 (9.7 to 20.7) | -5.7 (-11.0 to 0.5) | 0.72 (0.47 to 1.02) | 15.0 (9.9 to 21.0) | -5.2 (-10.7 to 1.0) | 0.74 (0.48 to 1.05) |
| Death | A | 13.3 (11.0 to 15.7) | -1.6 (-4.9 to 1.6) | 0.89 (0.71 to 1.11) | 13.2 (10.9 to 15.6) | -1.6 (-4.9 to 1.6) | 0.89 (0.71 to 1.12) |
|  | AB | 16.1 (8.5 to 23.8) | 1.2 (-6.6 to 8.9) | 1.08 (0.58 to 1.62) | 16.2 (8.7 to 23.5) | 1.4 (-6.4 to 8.9) | 1.10 (0.59 to 1.64) |
|  | B | 11.8 (8.6 to 15.0) | -3.1 (-7.0 to 0.6) | 0.79 (0.56 to 1.05) | 12.2 (9.0 to 15.5) | -2.6 (-6.6 to 1.3) | 0.83 (0.58 to 1.09) |
|  | O | 14.9 (12.9 to 17.1) | — | — | 14.8 (12.7 to 16.9) | — | — |
|  | Rh-pos | 14.5 (13.0 to 16.0) | — | — | 14.5 (13.0 to 16.0) | — | — |
|  | Rh-neg | 6.3 (3.0 to 10.1) | -8.2 (-11.7 to -3.8) | 0.44 (0.21 to 0.72) | 6.4 (3.0 to 10.3) | -8.2 (-11.7 to -3.7) | 0.44 (0.21 to 0.74) |

**Figure 1:**
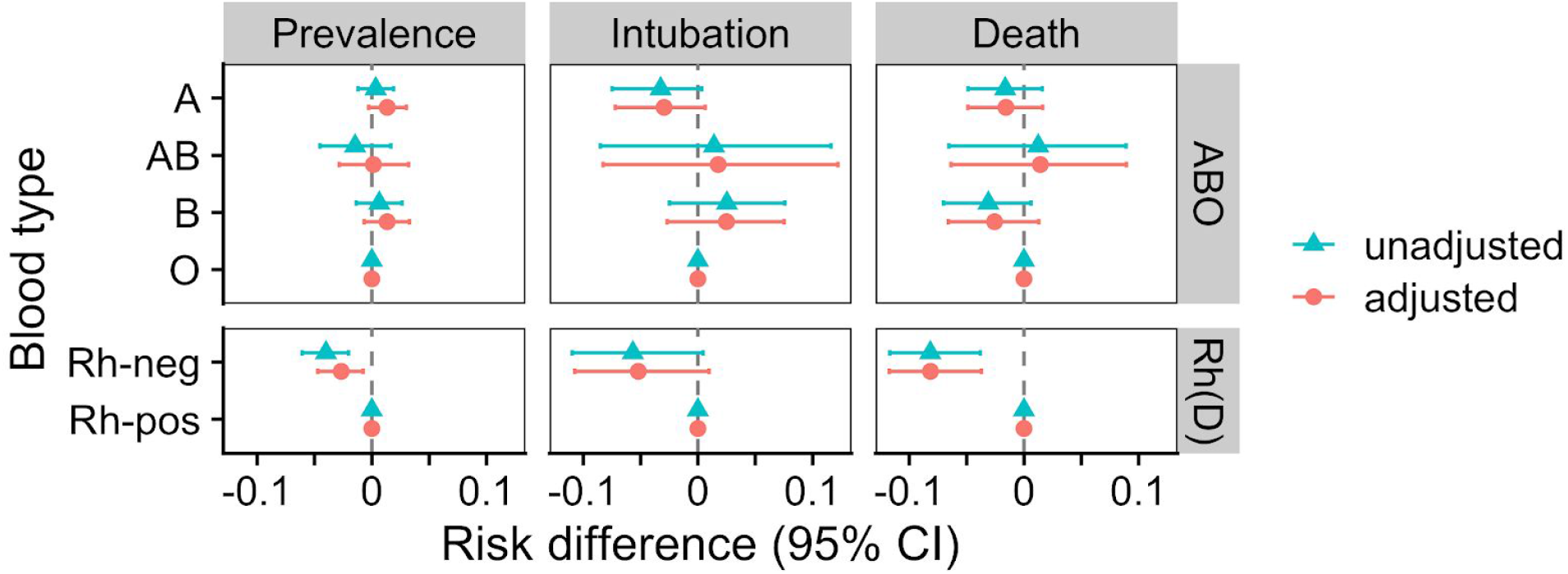
Estimated risk differences for blood types during the period from March 10 to August 1, 2020. Values represent risk differences for each blood type relative to the reference groups: O for ABO and positive for Rh(D). Prevalence risk differences were computed using linear regression, while intubation and death were computed using the Fine-Gray model [58]. Differences and 95% confidence intervals (CI) computed using Austin’s method, including bootstrap [54]. Adjusted models include race and ethnicity as covariates.

Blood type frequencies vary across ancestry groups [53], so we evaluated the confounding effect of ancestry by adjusting for race/ethnicity (proxies for ancestry). We compared infection prevalence with linear regression, using reference groups O for ABO and Rh-positive for Rh(D), and using bootstrap to compute 95% confidence intervals for each estimate [54]. With adjustment for patient race and ethnicity, prevalences among types A, AB, and B were higher than type O. Rh(D) negative individuals had a 2.7% lower risk of initial infection after adjustment for ancestry, consistent with a lower risk before adjustment.

Next, we examined intubation and death using a survival framework to understand how blood type affects progression to disease outcomes over time. Specifically, we used the Fine-Gray model [58] to estimate cumulative incidence functions by blood type while accounting for competing risks and adjusting covariates. Death and recovery were competing events for intubation and recovery was a competing event for death. Cohort entry was defined as the time of a patient’s first positive test result or the start of a hospital encounter if the first positive test occurred during the first 96 hours of the hospitalization. In accordance with CDC guidelines for returning to work [59,60], we defined a patient as having recovered only when 10 days had passed since the patient entered the cohort and only once the patient had been discharged. Patients appearing on before July 30 were considered, and outcomes beyond August 1, 2020 were censored.

Blood type A was at decreased risk of both intubation and death relative to type O, while type AB was at increased risk of both outcomes (Figure 1, Table 2). Conversely, we found that type B individuals were at higher risk of intubation but at lower risk of death, compared with type O. Individuals negative for Rh(D) were at decreased risk for both intubation and death, consistent with a lower risk of initial infection. Overall, we estimate between 0.1 and 8.2 percent absolute risk differences between blood groups, after adjusting for race and ethnicity.

## Discussion

Better understanding COVID-19 is imperative given the current pandemic’s toll. We investigated whether blood type is relevant for risk of infection, intubation, and death. Overall, we found modest but consistent risk differences between blood types. After adjusting for ancestry (the relevant confounder for this analysis), estimated risk differences were larger for intubation and death outcomes than for initial infection. We estimate larger risk differences between Rh blood types than between ABO types, with Rh-negative individuals being at lower risk of all three outcomes. Type A had lower risk of intubation and death compared with types AB and O. Only type B had inconsistent effects between intubation and death—type B increased risk of intubation and decreased risk of death compared to type O. We also found consistent evidence for protective associations between Rh negative blood groups and SARS-CoV-2 infection, intubation, death. Overall, blood type appears to have a consistent effect, though the magnitudes of these effects on risk of intubation or death are modest, and our estimates have large uncertainties relative to their magnitudes. The relatively large estimated errors in our analysis also suggest modest effect sizes and that greater sample sizes or meta-analyses are needed to estimate these effects more precisely.

After adjusting for ancestry by proxies of race and ethnicity, we found that types A and B conferred greater risk of an initial positive test compared to type O, while type AB (the rarest), conferred a very small risk decrease (0.2%). These results are consistent with an association discovered for SARS-CoV-1, in which O blood groups were less common among SARS patients [42]. Our results are also mostly consistent with the results reported by Zhao et al. [4], where non-O types appear to be at greater risk of infection, and with Ellinghaus et al. [5], where non-O appear to be at greater risk of infection but at lesser risk of mechanical ventilation, though the authors note that this decreased risk is not statistically significant at the 5% level. Unlike Ellinghaus et al., though, we estimate slightly higher risk for types B and AB relative to O for intubation.

Our results are based on data collected as part of hospital care during the early course of the pandemic, where outpatient testing was severely limited due to testing capacity and supply limitations. As such, our data are highly enriched for severely-ill patients, and the absolute risk values we report are not generalizable to all SARS-CoV-2-infected individuals. A considerable fraction of infections are mild or asymptomatic [61,62,63,64], while our data represent predominantly the most severe cases. Selection bias is a fundamental limitation of our study, so all our effect estimates are conditional on presentation to the hospital. Nonetheless, we minimized additional selection bias by making cohort criteria for cases and controls differ only with respect to the outcome of interest. Moreover, we found concordance between SARS-CoV-2-tested individuals and the general population at NYP/CUIMC in terms of blood type (Supplementary table 1). Consequently, our results are not affected by selection bias with respect to blood type, unlike some other blood type case/control study designs—particularly those using blood donors as controls, where enrichment of type O can be expected [5].

False negatives and time delay between test administrations and the return of their results both introduce noise to this analysis. We attempted to account for these biases by setting cohort entry at the time of first contact with the hospital when the patient tested positive less than 96 hours thereafter. This definition is imperfect, as 96 hours is sufficient for an individual infected shortly after admission to test positive (albeit with probability roughly 0.33) [65], but it is necessary to allow sufficient adjustment for the considerable time delay and retesting following false negatives. Another source of noise is the fact that not all intubations and deaths following a confirmed infection are related to COVID-19 (e.g. intubation during unrelated surgery). We defined recovery in an attempt to minimize this issue, though we recognize our definition is imperfect. Patients may be discharged prematurely and later return following onset of severe symptoms. Moreover, our 10 day cutoff for recovery is based on CDC guidelines for returning to work [59,60], which may be refined as additional evidence becomes available. Further work is needed to refine the definition of recovery and to determine which outcomes may be causally linked to COVID-19.

The *ABO* gene is highly polymorphic [66], and blood types have considerably different distributions across ancestry groups [53]. Like ABO, Rh groups are not distributed equally across race/ethnicity groups, with enrichment of Rh-negative among white and non-Hispanic individuals (Table 1). In addition, negative Rh blood groups are less common, representing only 9% of individuals in our data. While genetic data were not available for the patients included in our study, we used self-reported race and ethnicity as imperfect proxies for genetic ancestry. Adjusting for these covariates had a noticeable effect on our comparison of infection prevalence, but did not have an equally relevant effect on intubation or death (Figure 1, Table 2). This suggests that blood type may have a lesser, more confounded effect on infection prevalence than on intubation or death following confirmed infection. Nonetheless, race and ethnicity cannot fully capture ancestry, so the associations between blood types and COVID-19 we report may still be confounded by ancestry, even after adjustment. Further work is needed to better understand any potential residual confounding due to ancestry, not captured by race and ethnicity.

## Conclusion

In this study we found evidence for associations between ABO and Rh blood groups and COVID-19. Using data from NYP/CUIMC, we found moderately increased infection prevalence among non-O blood types and among Rh-positive individuals. Intubation risk was increased among AB and B types, and decreased among A and Rh-negative types. Risk of death was slightly increased among type AB individuals and was decreased among types A, B, and Rh-negative types. All estimates were adjusted for patient ancestry using self-reported race and ethnicity. Our results add further evidence to the previously discovered associations between blood types and COVID-19.

## Data Availability

NYP/CUIMC Patient data is protected by HIPAA and cannot be released. However, we are releasing summary information and data from analyses.

https://github.com/zietzm/abo_covid_analysis

## Acknowledgements

We would like to thank Ben May and Vijendra Ramlall for assistance with data collection and periodic updates to the patient data. We would also like to thank Nicholas Giangreco, Undina Gisladottir, and Dr. Phyllis Thangaraj for helpful discussions regarding risk factor definitions. MZ is funded by NIH T15 LM007079, and NPT is funded by R35GM131905.

## Author contributions

MZ and NPT conceived and designed the study. MZ carried out the statistical analysis with advice from NPT. JZ and MZ created the cohort entry and exit criteria. MZ and NPT wrote, revised, and approved the final version of the manuscript.

## Methods

We identified the cohort for this study by filtering the NYP/CUIMC data warehouse for patients with a recorded SARS-CoV-2 test and those having a recorded blood type. Next, we removed any individual with multiple, contradictory blood type measurements, reflecting likely errors in the data. Finally, we excluded individuals below age 18 from our analysis.

Blood group was determined using laboratory measurements coded using descendant concepts of LOINC LP36683-8 (ABO and Rh group). Individuals with contradictory measurements were excluded. Intubation was assessed using completed procedures having the procedure description, “Intubation.” We grouped race into five categories and ethnicity into two. Specifically, we considered only Asian, Black/African-American, and White, categorizing other listed races (all of which were small minorities) as ‘Other’, and missing or declined race as ‘Missing’. Ethnicity was grouped as either Hispanic or non-Hispanic.

We sought to estimate total effects of blood type on COVID-19 outcomes. Using a graphical model (Supplementary Figure S2), we identified ancestry as the only confounding variable for an estimate of total effect, since blood type is genetic and varies across ancestry groups. As genetic data were not available, we used self-reported race and ethnicity as proxies for ancestry. We were unable to identify a method to alleviate selection bias in our data, so the effects we report are conditional on presence at NYP/CUIMC.

We considered three outcomes: initial infection, intubation, and death. Our evaluation of initial infection sought to assess the infection prevalence differences among individuals presenting to the hospital, not those potentially infected at the hospital or long after their first test. Due to the high risk for false negatives [55,56], we considered any positive test less than 96 hours after the start of an encounter as evidence of initial infection. Initial infection risk differences between blood types were assessed using linear regression, and race/ethnicity were adjusted by including them as covariates. We used Austin’s bootstrap method to compute 95% confidence intervals for all risk estimates [54], using 1000 bootstrap iterations.

We assessed intubation and death as severe outcomes of COVID-19, and evaluated blood type effects using survival analysis. Individuals entered the at-risk cohort either at the time of their first positive test result, or at the time of first contact with the hospital when the first positive test occurred within 96 hours of the start of a hospital encounter. Patients with Do-Not-Intubate orders were excluded from consideration for the intubation outcome. We defined a patient as recovered only after being discharged from the hospital and only once 10 days have passed since cohort entry. Death and recovery are competing risks for intubation, and recovery is a competing risk for death. Finally, outcomes beyond August 1 were censored, as this was the last date for which we have data available. Intubation and death were assessed using Fine-Gray models, which can estimate cumulative incidences. As before, race and ethnicity were adjusted by including them as covariates, and confidence intervals were computed with 1000 bootstrap iterations.

This study is approved by the IRB (#AAAL0601). We conducted our analyses using the R language, using the cmprsk package [67] implementation of the Fine-Gray model. The manuscript was written openly on GitHub using Manubot [68].

## Data availability

While our data from NYP/CUIMC are protected by HIPAA and cannot be released, we have made all code used for our analysis available at https://github.com/zietzm/abo_covid_analysis.

## Code availability

We have made all code used for our analysis available at https://github.com/zietzm/abo_covid_analysis.

## Supplemental information

**Supplementary Figure 1:**
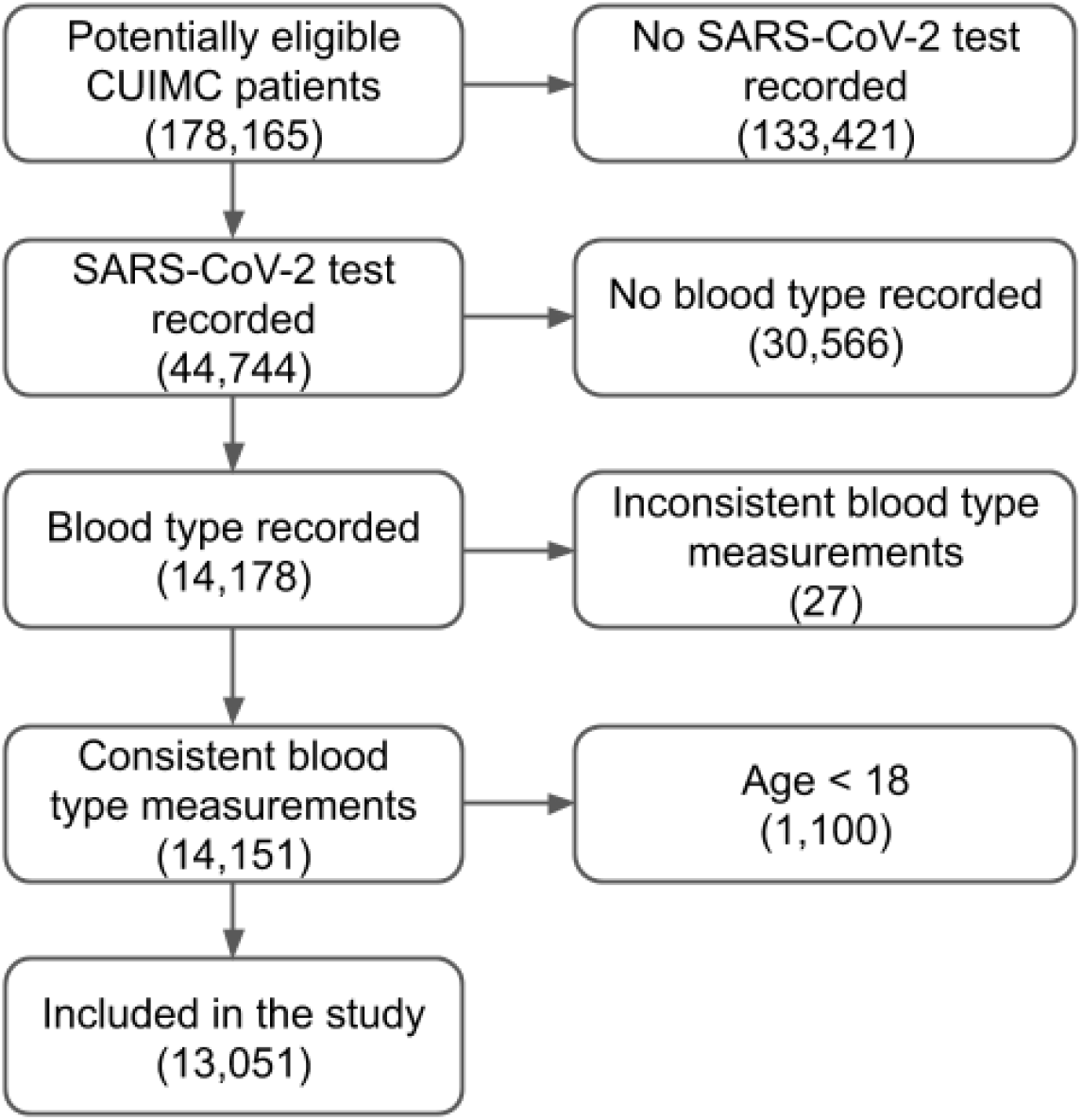
Flow diagram of inclusion and exclusion criteria for the cohort used. Numbers indicate the number of patients in each group. Groups on the right were excluded.

**Supplementary Figure 2:**
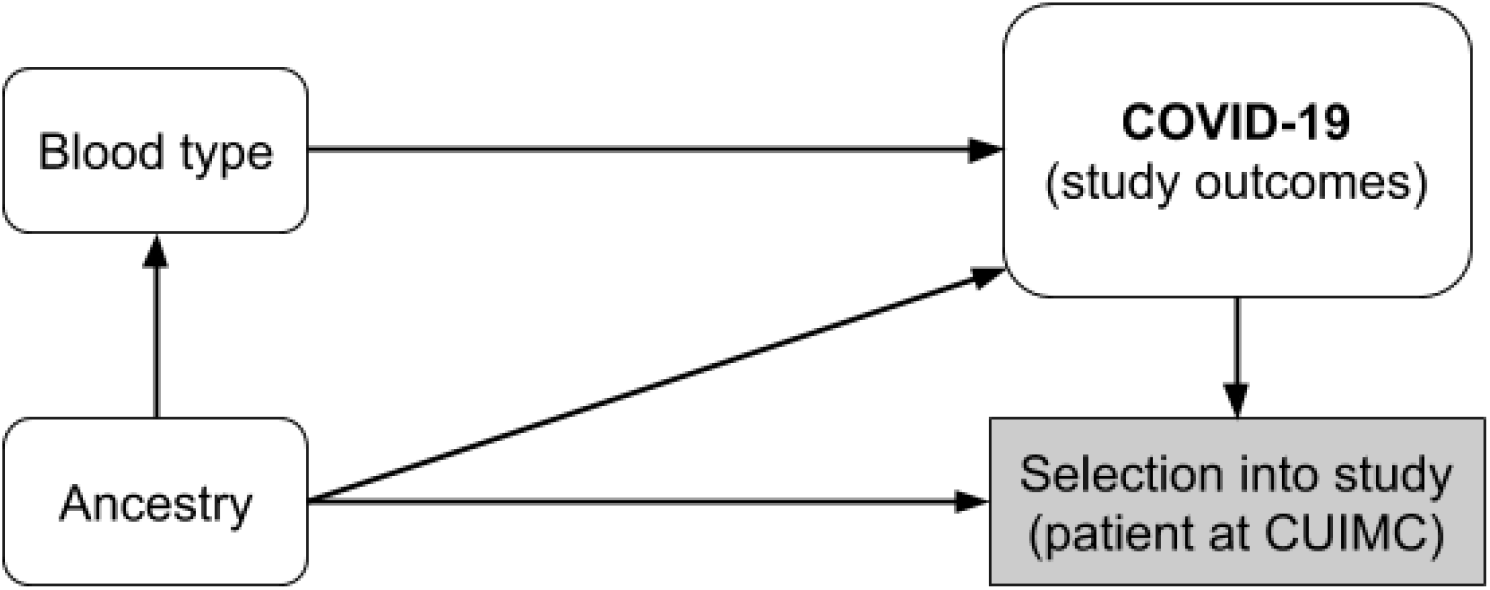
Graphical model of the system under investigation. We sought to estimate the total effects of blood type on the COVID-19 outcomes under investigation. Confounding can be controlled by adjusting for ancestry. Selection bias cannot be controlled fully, and as a result, our estimates are conditional on presentation to the hospital during the COVID-19 pandemic.

**Supplementary Table 1:** Chi-squared tests to evaluate whether the dependence between blood type and having received a test for SARS-CoV-2. ABO had three degrees of freedom, while Rh(D) had one degree of freedom.

| Blood group | SARS-CoV-2 tested | non-SARS-CoV-2-tested | Chi-squared | p-value |
| --- | --- | --- | --- | --- |
| ABO | A: 4298 (32.9%), AB: 559 (4.3%), B: 2033 (15.6%), O: 6161 (47.2%) | A: 34156 (32.7%), AB: 4405 (4.2%), B: 15590 (14.9%), O: 50305 (48.2%) | 5.79 | 0.122 |
| Rh(D) | neg: 1195 (9.2%), pos: 11856 (90.8%) | neg: 9644 (9.2%), pos: 94812 (90.8%) | 0.0716 | 0.789 |

